# Aerobic fitness is associated with cerebral mu-opioid receptor availability and activation in healthy humans

**DOI:** 10.1101/2020.12.13.20247627

**Authors:** Tiina Saanijoki, Tatu Kantonen, Laura Pekkarinen, Kari Kalliokoski, Jussi Hirvonen, Lauri Tuominen, Jetro J. Tuulari, Eveliina Arponen, Pirjo Nuutila, Lauri Nummenmaa

## Abstract

Central μ-opioid receptors (MORs) modulate affective responses to physical exercise. Individuals with higher aerobic fitness report greater exercise-induced mood improvements than those with lower fitness, but the link between cardiorespiratory fitness and the MOR system remains unresolved. Here we tested whether maximal oxygen uptake (VO_2peak_) and physical activity level are associated with cerebral MOR availability, and whether these phenotypes predict endogenous opioid release following aerobic exercise. We studied 64 healthy lean men who performed a maximal incremental cycling test for VO_2peak_ determination, completed a questionnaire assessing moderate-to-vigorous physical activity (MVPA, min/week), and underwent positron emission tomography with [^11^C]carfentanil, a specific radioligand for MOR. A subset of 24 subjects underwent additional PET scan also after a one-hour session of moderate-intensity exercise. Higher VO_2peak_ and self-reported MVPA level was associated with larger decrease in cerebral MOR binding after aerobic exercise in ventral striatum, orbitofrontal cortex and insula. That is, higher fit and more trained individuals showed greater opioid release acutely following exercise in brain regions especially relevant for reward and cognitive processing. Higher VO_2peak_ also associated with lower baseline *BP*_ND_ in the reward and pain circuits, i.e., in frontal and cingulate cortices as well as in temporal lobes and subcortically in thalamus and putamen. We conclude that higher aerobic fitness and regular exercise training may induce neuroadaptation within the MOR system which might contribute to improved emotional and behavioural responses associated with long-term exercise.

## Introduction

Habitual physical activity and cardiorespiratory fitness (CRF) are well-established modifiable lifestyle factors that promote brain health throughout the lifespan. Higher fitness and greater amounts of physical activity are linked with better cognitive functioning [1,2], lower levels of anxiety and depression [3,4], and reduced risk for neurodegenerative disease [5]. These biological and psychological benefits of exercise are paralleled in brain structure and function. Better fitness and higher physical activity levels are associated with higher grey [6–8] and white matter volume [6,9,10]. In addition, several intervention studies have demonstrated that improved fitness positively affects brain volumes in older adults, especially in frontotemporal regions that are important for cognition and memory functions, and most susceptible to age-related brain atrophy [11–14]. Moreover, higher aerobic fitness promotes efficient functional connectivity of multiple brain networks supporting cognitive control and memory functions [15,16].

Physical exercise also acutely affects functioning of the brain’s neuromodulatory systems, particularly the endogenous opioid system [17]. Endogenous opioid system and especially μ-opioid receptors (MORs) are closely involved in processing reward [18], motivation [19,20], and emotions [21]. They also have a central role in several physiological functions such as pain processing [22] and stress regulation [23,24], and recent evidence links opioid system dysregulation with depressive and anxious symptoms [25]. Therefore, the opioid system could potentially mediate the psychological benefits of regular exercise such as improved mood.

Animal studies investigating the effects of regular exercise training on the opioid system have found elevated β-endorphin and met-enkephalin levels in periaqueductal grey area and rostral ventromedial medulla after five weeks of treadmill running [26] and shown that chronic exercise, in comparison with short-term exercise or no exercise, decreases MOR expression [27] and overall MOR availability in rat brain [28]. Exercising rats also show decreased sensitivity to antinociceptive effects of exogenous opioid agonists such as morphine, which may indicate downregulation of MORs resulting from increased endogenous opioid concentrations elevated by regular exercise training [29,30]. Human neuroimaging studies have demonstrated that a single bout of moderate-to-vigorous exercise stimulates endogenous opioid release in the brain, which is associated with affective responses induced by exercise [31–33]. Taken together, converging evidence from animal and human studies suggest that regular exercise training might induce neuroadaptation within central MOR system, subsequently contributing to improvements in mood and stress regulation. Yet, *in vivo* evidence from humans is currently lacking. Here we investigated whether individual differences in baseline MOR availability are associated with cardiorespiratory fitness and habitual physical activity levels in healthy young men. We used *in vivo* PET imaging with the highly selective MOR agonist ligand [^11^C]carfentanil. We coupled MOR data with measurement of VO_2peak_, an objective and direct measure of CRF, and with self-reported physical activity questionnaires. To test whether higher fitness and physical activity levels influence the capacity of acute exercise to activate the MOR system, we also studied a subset of participants with [^11^C]carfentanil PET after a one-hour session of aerobic exercise. Based on previous human and animal research, we hypothesized that higher levels of fitness and physical activity would be negatively associated with cerebral MOR availability in the brain’s reward circuits and positively associated with GM volume. We predicted that VO_2peak_ and self-reported physical activity would be associated with exercise-induced changes in MOR availability.

## Materials and Methods

### Subjects

The Ethics Committee of the Hospital District of Southwest Finland approved the study protocol, and the study was conducted in accordance with the Declaration of Helsinki. All subjects signed ethics-committee-approved informed consent forms. 64 male adults with a variable exercise background were enrolled in the study (Table 1). They were recruited via Internet discussion forums, traditional bulletin boards, university-hosted email lists and newspaper advertisements. The exclusion criteria were poor compliance, a history of or current neurological or psychiatric disease, use of tobacco products or medication affecting the central nervous system, current or past excessive alcohol or substance abuse, as well as standard PET and MRI exclusion criteria. Laboratory tests, urinalysis, and an ECG were obtained to assess health and the absence of psychoactive drugs. These data have originally been collected in clinical trials EXEBRAIN (NCT02615756) and PROSPECT (NCT03106688).

**Table 1.** Subject characteristics (n=64).

|  | Mean (SD) | Range |
| --- | --- | --- |
| Age (years) | 25.4 (4.6) | 20–36 |
| Body Mass Index ($\text{kg}/\text{m}^2$) | 24.1 (2.8) | 18.5–31.0 |
| Total physical activity (min/week) | 291 (165) | 0–870 |
| Peak oxygen consumption<br>($\text{VO}_{2\text{peak}}$; $\text{ml}/\text{kg}/\text{min}$) | 44.5 (7.9) | 25.8–61.7 |

### Physical activity and aerobic fitness measurements

Self-reported physical activity was assessed with a questionnaire where participants rated the frequency (days/week) and duration (hours and minutes/week) of moderate to vigorous physical activity (MVPA) and other physical activity during the last three months. CRF was assessed as peak oxygen consumption (VO_2peak_), which was determined in a maximal exercise test performed on a cycle ergometer starting at 40-50 W and followed by an increase of 30 W in every 2 minutes until volitional exhaustion. Ventilation and gas exchange were measured (Jaeger Oxycon Pro; VIASYS Healthcare) and reported as the mean value per minute. The highest 1-min mean value of oxygen consumption was expressed as the VO_2peak_.

### PET data acquisition

We measured MOR availability with the agonist radioligand [^11^C]carfentanil that has high affinity for MORs. Radioligand synthesis for the EXEBRAIN trial has been described previously [32]. For the scans from the PROSPECT trial, [^11^C]carfentanil was synthesized using [^11^C]methyl triflate, where cyclotron-produced [^11^C]methane was halogenated by gas phase reaction into [^11^C]methyl iodide [34] and converted online into [^11^C]methyl triflate [35]. The [^11^C]methane was produced at the Accelerator Laboratory of the Åbo Akademi University, using the ^14^N(p,α)^11^C nuclear reaction in a N_2_-H_2_ target gas (10 % H_2_). [^11^C]methyl triflate was bubbled into a solution containing acetone (200 µl), O-desmethyl precursor (0.3– 0.4 mg, 0.79–1.05 µmol) and tetrabutylammonium hydroxide (aq) (4 µl, 0.2 M) at 0 °C. The reaction mixture was diluted and loaded into a solid phase extraction cartridge (C18 Sep-Pak^®^ Light, Waters Corp., Milford, MA) and the cartridge was washed. Dilution and washing were done using 25% ethanol in sterile water solution, 10 mL each step. The [^11^C]carfentanil was extracted with ethanol from the cartridge, diluted with 0.1 M phosphate buffer solution into < 10 % ethanol level and finally sterile filtered (Millex GV, 0.22 µm polyvinylidene fluoride membrane, 33 mm, Merck Millipore). Analytical HPLC column (Phenomenex Luna^®^ 5 µm C8(2) 100 Å, 4.6 × 100 mm), acetonitrile (32.5%) in 50 mM H_3_PO_4_ mobile phase, 1 ml/min flow rate, 7 min run time and detectors in series for UV absorption (210 nm) and radioactivity were used for determination of identity, radiochemical purity and mass concentration. Radiochemical purity of the produced [^11^C]carfentanil batches was 98.5 ± 0.3 % (mean ± SD). The injected [^11^C]Carfentanil radioactivity was 248 ± 11 MBq and molar radioactivity at time of injection 290 ± 110 MBq/nmol corresponding to an injected mass of 0.40 ± 0.23 µg.

Subjects refrained from exercise at least 24 hours and fasted for at least 2 h before scanning. Data were acquired with the 3T Philips Ingenuity TF PET/MR (PhilipsHealthcare, Cleveland, OH, USA) scanner or PET/CT (GE Discovery VCT PET/CT, GE Healthcare (General Electric Medical Systems, Milwaukee, WI, USA) at Turku PET Centre. Data acquisition started concomitantly with the intravenous radioligand bolus-injection (M = 250 MBq, SD = 13 MBq), and cerebral radioactivity was measured for 51 min. Data were corrected for dead-time, decay, and measured photon attenuation.

### PET challenge paradigm for exercise-induced opioid release

A subset of participants (n = 23) underwent an additional PET scan after a one-hour session of moderate-intensity cycling exercise on a separate day; the protocol and opioid release data have been reported previously [32]. The order of the exercise / rest PET studies was randomized and counterbalanced for these participants. Emotional reactions to physical exercise were measured with Positive Affect and Negative Affect Schedule [36].

### MRI acquisition

Anatomical MR images were acquired for VBM as well as for preprocessing the PET images with the 3T Philips Ingenuity TF PET/MR scanner using a T1-weighted sequence with 1 mm^3^ resolution (TR 8.1 ms, TE 3.7 ms, flip angle 7°, scan time 263 s). Complementary voxel-based morphometric analyses on the association between aerobic fitness, physical activity and cerebral density are described in SI.

### PET data preprocessing and analysis

PET data were processed with the automated Magia pipeline [37] (https://github.com/tkkarjal/magia). Processing began with motion-correction of the PET data followed by coregistration of the PET and MR images. Magia uses FreeSurfer (http://surfer.nmr.mgh.harvard.edu/) to define the regions of interest (ROIs) as well as the reference regions. The ROI-wise kinetic modeling was based on extraction of ROI-wise time-activity curves. The PET images were slightly smoothed using Gaussian kernel (2 mm full width at half maximum, FWHM) to increase signal-to-noise ratio before model fitting Parametric images were spatially normalized to MNI-space and finally smoothed using a Gaussian kernel (FWHM = 6 mm). [^11^C]carfentanil binding was quantified by binding potential (*BP*_ND_), which is the ratio of specific binding to non-displaceable binding in the tissue [38]. Occipital cortex was used as the reference region [39].

### Statistical analysis

The effects of VO_2peak_ and self-reported physical activity on i) MOR availability, ii) MOR activity following physical exercise, and iii) on GM densities were assessed in SPM12 (http://www.fil.ion.ucl.ac.uk/spm/) using linear regression model with BMI and PET scanner (for PET data) as a covariate. Statistical threshold was set at *p* < 0.05, FDR-corrected at cluster level. Atlas-based ROIs were generated in the MOR-rich regions in the brain (amygdala, hippocampus, ventral striatum, dorsal caudate, thalamus, insula, orbitofrontal cortex (OFC), anterior cingulate cortex (ACC), middle cingulate cortex (MCC), and posterior cingulate cortex (PCC) using AAL [40] and Anatomy [41] toolboxes. Mean regional [^11^C]carfentanil *BP*_ND_ were extracted for each region, and the averaged ROI data were analysed with R statistical software (https://cran.r-project.org) [42].

## Results

### VO_2peak_ is associated with physical activity level, age, and BMI

VO_2peak_ was positively associated with self-reported physical activity (r = 0.45, *p* < 0.01) and negatively associated with age (r = -0.37, *p* < 0.01) and BMI (r = -0.46, *p* < 0.01). Age was positively associated with BMI (r = 0.38, p < 0.01).

### Higher VO_2peak_ and MVPA level predict larger decrease in MOR availability after exercise

There were no significant differences in [^11^C]carfentanil *BP*_ND_ between baseline and moderate-intensity exercise conditions at group level, as reported in previously published work [32]. However, change in MOR availability varied notably between individuals following aerobic exercise, such that *BP*_ND_ decreased in some individuals but increased in others. We then tested whether exercise-induced changes in *BP*_ND_ would be associated with self-reported physical activity or VO_2peak_, indicative of exercise habit-dependent MOR activation. We found a negative association between VO_2peak_ and exercise-induced change in *BP*_ND_, such that higher VO_2peak_ was associated with larger decrease in *BP*_ND_ after exercise (**Fig. 1**). This effect was observed in ventral and dorsal striatum, left hippocampus, left thalamus, insular cortex, somatosensory cortex, temporal areas, and orbitofrontal cortex. Exercise-induced change in *BP*_ND_ was also correlated with self-reported MVPA (**Fig. 2**), however no associations were found between total self-reported physical activity and exercise-induced change in *BP*_ND_.

**Figure 1.**
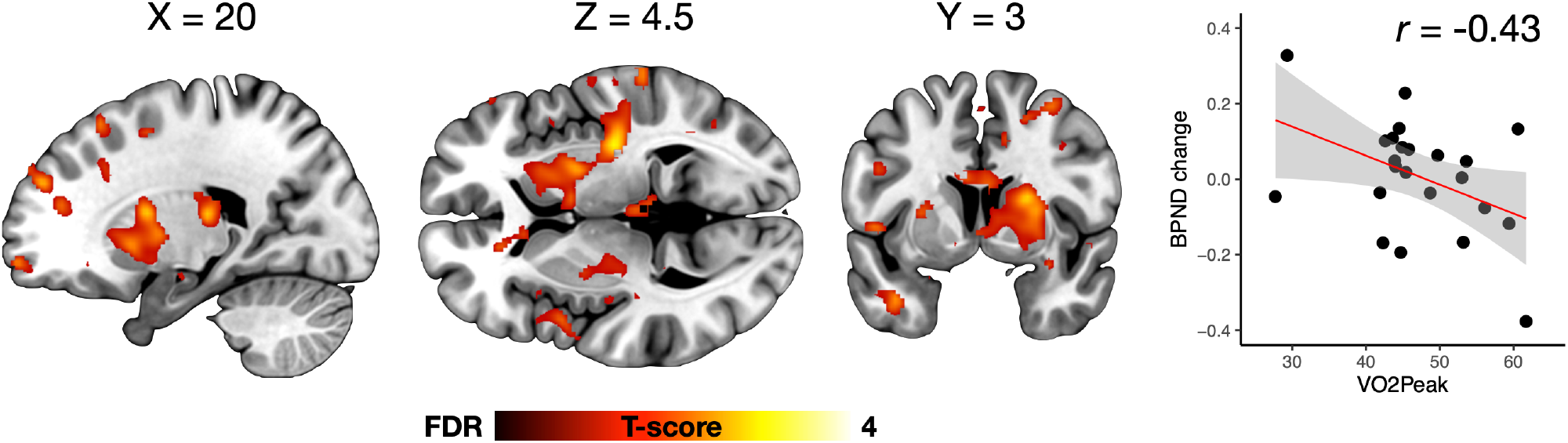
Higher aerobic fitness predicted higher exercise-induced opioid release after moderate-intensity exercise, as indicated by a negative association between VO_2peak_ and change in BP_ND_ after one-hour session of aerobic exercise. The data are thresholded at p < 0.05, FDR-corrected at the cluster level. Scatterplot shows the corresponding association (LS-regression line with 95% CI) in putamen.

**Figure 2.**
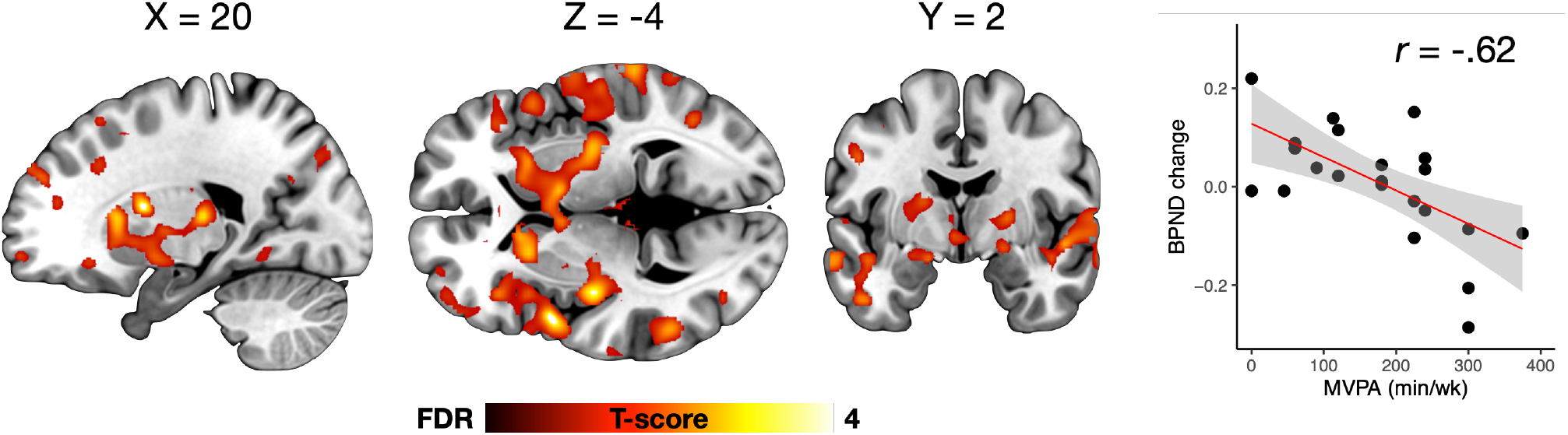
Higher training level predicted higher exercise-induced opioid release after moderate-intensity exercise, as indicated by a negative association between self-reported moderate-to-vigorous physical activity (MVPA) level and changes in BP_ND_ after one-hour session of aerobic exercise. The data are thresholded at p < 0.05, FDR-corrected at the cluster level. Scatterplot shows the corresponding association (LS-regression line with 95% CI) in the right fusiform gyrus.

We previously reported enhanced mood responses after aerobic exercise [32]. Here, we found a positive association between VO_2peak_ and change in positive affect as measured with Positive Affect and Negative Affect Schedule before and after aerobic exercise (r = 0.59, *p* < 0.01), indicating that higher VO_2peak_ was associated with higher mood improvement.

### Fitness and physical activity level are negatively associated with baseline MOR availability

We next tested whether baseline differences in aerobic fitness are associated with MOR availability. Full-volume analysis showed a negative association between VO_2peak_ and baseline MOR availability (*BP*_ND_) in a large cluster extending to both hemispheres from the frontal lobe to the parieto-occipital sulcus (**Fig. 3**). Significant associations were also observed in bilateral putamen, thalamus, insula, and temporal cortices. The ROI analysis revealed significant associations in orbitofrontal and middle cingulate cortices (*p*s < 0.05). Comparable analysis where MOR availability was predicted with self-reported physical activity yielded similar effects, but only when BMI was not controlled for in the model (data not shown).

**Figure 3.**
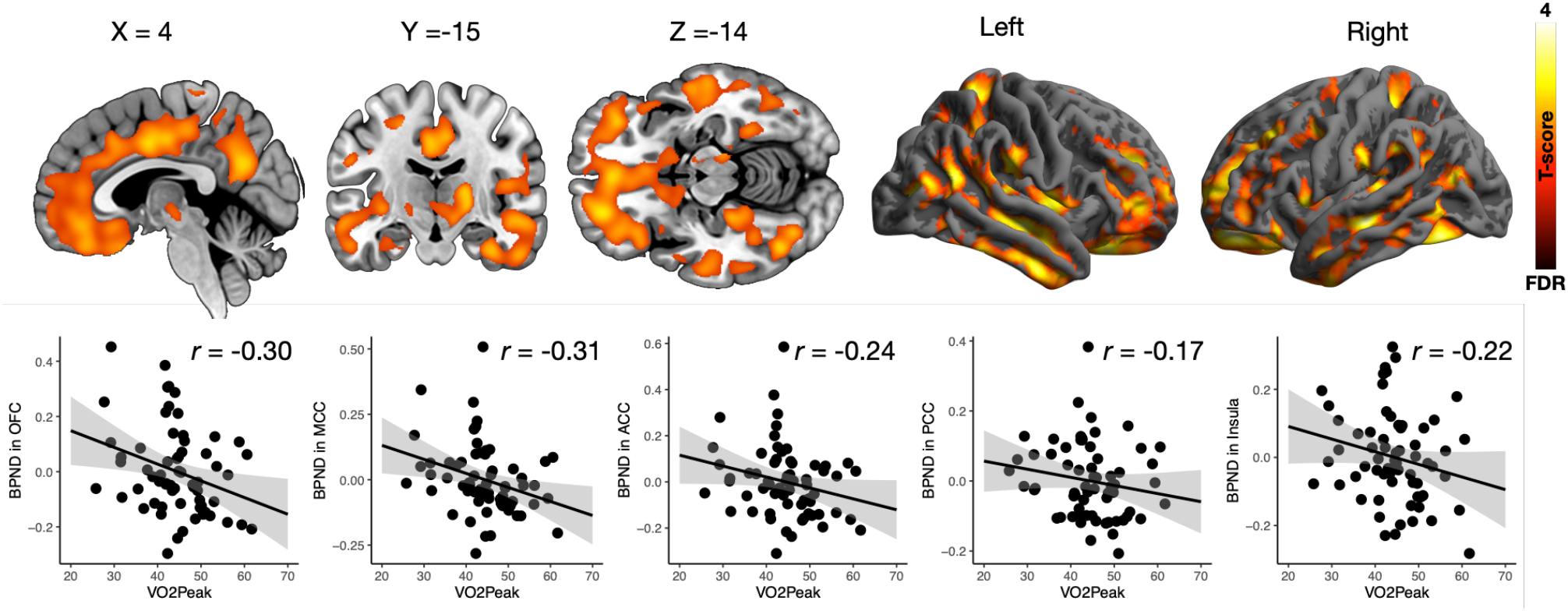
Negative association between VO_2peak_ and [^11^C]carfentanil BP_ND_. The data are thresholded at p < 0.05, FDR-corrected at the cluster level. Scatterplots show the corresponding association (LS-regression line with 95% CI) in representative anatomical regions of interest.

Ageing has regionally specific effects on MOR availability [43]. Although our subjects had a relatively narrow age range (M = 25.4, SD = 4.6) we nevertheless wanted to statistically control for potential ageing-dependent effects in the MOR availability. When age was entered in the analysis as a covariate, no associations were observed between MOR availability and VO_2peak_ or self-reported physical activity at the a priori statistical threshold. However, with more lenient thresholding (p < 0.05 uncorrected) the spatial pattern of results remained similar as shown in **Fig 3**. VO_2peak_ showed positive association with GM density when controlling for age and BMI (Supplementary Figure S1). These findings are described in detail in Supplementary Information.

## Discussion

The present findings indicate that aerobic fitness has a previously unrecognized role in brain opioid signaling. Higher cardiorespiratory fitness, as measured by VO_2peak_, and higher self-reported moderate-to-vigorous physical activity (MVPA) level, were associated with larger decrease in cerebral MOR binding after aerobic exercise in ventral striatum, orbitofrontal cortex and insula. In other words, higher-fit individuals showed greater acute opioid release following exercise in brain regions involved in reward and cognitive processing. We also found that higher VO_2peak_ was associated with lower baseline *BP*_ND_ in the reward and pain circuits, i.e., in frontal and cingulate cortices as well as in temporal lobes and subcortically in thalamus and putamen. Conversely, VO_2peak_ was positively associated with GM density in several brain regions including medial and lateral frontal and orbitofrontal cortices, cingulate cortex and striatum. Taken together, these data suggest that aerobic fitness may modulate cerebral MOR tone and function which might be an important pathway regulating exercise habits, and that aerobic fitness and physically active lifestyle protect against GM loss not only in older adults but also in early adulthood.

### Higher aerobic fitness predicts greater decrease in MOR binding after aerobic exercise

Previous research has shown that exercise intensity modulates opioid action in the brain [32,33]. High-intensity exercise induces a robust opioid release, whereas moderate-intensity exercise results in decreased MOR availability in some and increased in other individuals [32]. Here we report, for the first time, that both higher VO_2peak_ and MVPA levels are associated with higher cerebral opioid release after a bout of moderate-intensity exercise, suggesting that aerobic fitness and physical activity level may shape opioidergic response following aerobic exercise. This dependency on prior aerobic fitness likely explains why moderate-intensity exercise does not result in net opioid release when individuals with different fitness status are assessed together [32]. The present findings go beyond past reports, which have only examined the association of training status on peripheral opioid concentrations after exercise. Higher circulating β-endorphin levels has been found in well-trained athletes, in comparison with untrained individuals, after a graded exercise test [44] and after a bout of supramaximal exercise [45]. In contrast, high-intensity cycling (70% and 80% of VO_2max_) resulted in similar increase in plasma β-endorphin concentration in both trained and untrained individuals whereas moderate-intensity cycling (60% of VO_2max_) showed no effect on plasma β-endorphin concentration [46]. While it has been suggested that training-induced adaptation within the opioid system could increase the response capacity to extreme exercise stress [45], peripheral opioid levels probably do not mirror those of the brain [47] and thus limits reasonable comparison of these studies.

We observed greater opioid release in higher fit participants after aerobic exercise in ventral and dorsal striatum, left hippocampus, left thalamus, insular cortex, somatosensory cortex, temporal areas, and orbitofrontal cortex. MORs in these regions are closely involved in processing both nociceptive and hedonic signals [21,48] as well as modulating decision making and cognitive control [49]. We found that higher fit participants experienced greater improvements in mood following the aerobic exercise. This accords with previous studies reporting that better fitness level [50] and regular exercise participation are associated with more positive affective responses [51–53] and enhanced anxiety relief [52,54] following a bout of exercise. Antinociceptive effects of acute exercise have also been found to depend on fitness level [55]. We propose that greater opioid release could explain enhanced emotional and antinociceptive responses reported by people with higher exercise levels and thus, bear implications in long-term exercise engagement.

### Greater aerobic fitness and physical activity level are associated with lower MOR availability

We found that VO_2peak_ and self-reported physical activity were negatively correlated with baseline [^11^C]carfentanil *BP*_ND_, suggesting that higher levels of regular exercise promoting better aerobic fitness may induce neuroadaptation within the endogenous opioid system. This accords with prior animal studies that have established a relationship between habitual physical activity and endogenous opioid system [27–30]. Chronic exercise, in comparison with short-term exercise or no exercise, decreases MOR expression [27] and overall MOR binding in rat brain [28], demonstrating a causal link between exercise and MOR binding. We observed associations between VO_2peak_ and *BP*_ND_ mainly in cortical regions including prefrontal, cingulate, insular and parahippocampal cortices, but also in subcortical regions, such as putamen and thalamus. As these regions are closely implicated in the modulation of emotions [21,48] and cognitive processes [49], functional changes in these regions may mediate many psychological benefits derived from regular exercise practice as well as exercise motivation and habits. In line with this, functional imaging studies have consistently reported associations between various indices of fitness such as VO_2peak_ or blood lactate curve and executive functions in lateral fronto-parietal and anterior cingulate cortices [56–60]. Additionally, temporal regions have been suggested to play a role in regular exercise induced mood improvements [61], and insula and parietal cortex have shown altered reward processing in response to regular exercise training [62]. Given the found association between aerobic fitness and MOR availability in these regions, enhanced opioid modulation following improved fitness might contribute to long-term exercise-induced benefits in emotional and cognitive processing.

Reduced MOR binding may reflect either down-regulation of MORs, increased opioid tone and following competition between endogenous opioids and the radioligand, or a combination of the two. Previous animal studies suggest that regular exercise training increases tonic endogenous opioid levels. In rats, exercise training for five to eight weeks increases basal β-endorphin concentration in cerebrospinal fluid [63] and plasma [64] and elevates both β-endorphin and met-enkephalin levels in periaqueductal grey area and rostral ventromedial medulla [26]. Such rise in tonic opioid levels is also associated with altered pain processing, as regular exercise training increases nociceptive threshold in rats [65], reverses measures of pain in animal models of chronic pain [26,66,67], and as these effects can be reversed by an opioid receptor antagonist naloxone [26,65–67]. Sensitivity to antinociceptive effects of exogenous opioid agonists such as morphine also decreases in exercising animals, suggesting that chronic exercise induces cross-tolerance to exogenously administered opioid agonists due to greater concentrations of endogenous opioid peptides [29,30]. Higher opioid levels may also contribute to long-term adaptations of other physiological and behavioural responses associated with regular exercise, such as improved mood and stress regulation, but may also be implicated in exercise addiction [30].

Altogether our findings suggest that improving aerobic fitness by regular physical activity of moderate to high intensity could influence MOR system in two ways. First, by altering the baseline MOR availability and second, by improving exercise-induced opioid functioning by enhancing the capacity of single session of aerobic exercise to activate the MOR system and concomitant positive affective responses. Such exercise training derived MOR adaptation might further facilitate long-term exercise motivation and continuous exercise practice [68,69], yet this idea remains to be determined in future studies.

### Limitations

In the present study, both age and BMI correlated positively with MOR availability. However, prior work with larger sample has shown that ageing increases MOR availability in frontotemporal areas and decreases it in thalamus and nucleus accumbens, and that MOR availability has no significant association with BMI [43]. Thus, pure age effects would unlikely explain our findings in the thalamus. Despite the narrow age range (20-36 years) of our subjects, when age was included as a covariate in the model between baseline MOR availability and aerobic fitness or self-reported physical activity, the effects were statistically less significant. Consequently, it is difficult to disentangle the presently observed fitness-dependent effects on baseline MOR availability from the joint effects between fitness and age.

## Conclusions

We conclude that higher aerobic fitness and higher MVPA level are associated with greater reductions in MOR availability after a bout of aerobic exercise, suggesting greater exercise-induced opioid release in high fit and more trained individuals. Moreover, aerobic fitness was negatively associated with baseline MOR availability and positively associated with grey matter density. Lower MOR binding in higher fit participants in the present study may result from higher endogenous opioid levels following regular exercise training, yet this remains speculative as the outcome measure *BP*_ND_ does not distinguish between receptor density, affinity, and the amount of endogenous neurotransmitter occupancy. Nevertheless, the associations between VO_2peak_ and baseline MOR availability and reduced MOR availability after exercise suggest that regular exercise training may induce neuroadaptation within the MOR system also in humans, which may further modulate physiological and behavioural responses governed by the opioid system.

## Data Availability

Per Finnish legislation, the medical images are considered as sensitive personal data and may thus not be distributed.

## Funding

This study was supported by the Academy of Finland (grants #251125, #251399, #256470, #265917, #281440, #283319, #304385, and #332225) and Sigrid Juselius foundation. We thank Emil Aaltonen Foundation, Finnish Cultural Foundation (Southwest Finland Fund), and Jenny and Antti Wihuri Foundation for personal grants to TK. The authors declare no conflict of interest.

Supplementary information accompanies this paper.

## Notes

### Competing Interest Statement

The authors have declared no competing interest.

### Clinical Trial

NCT02615756, NCT03106688

### Author Declarations

The Ethics Committee of the Hospital District of Southwest Finland

