## Supplemental information for "Aerobic fitness is associated with cerebral mu-opioid receptor availability and activation in healthy humans"

1) Turku PET Centre, University of Turku, Turku, Finland, 2) Turku BioImaging, University of Turku and Åbo Akademi University, Turku, Finland, 3) Clinical Neurosciences, University of Turku and Turku University Hospital, Turku, Finland 4) Department of endocrinology, Turku University Hospital, Turku, Finland, 5) Department of Radiology, Turku University Hospital, Turku, Finland, 6) The Royal's Institute of Mental Health Research, University of Ottawa, Ottawa, ON, Canada, 7) FinnBrain Birth Cohort Study, Turku Brain and Mind Center, Department of Clinical Medicine, University of Turku, Turku, Finland, 8) Department of Psychology, University of Turku, Turku, Finland

### Supplementary methods

#### Voxel-based morphometry

Voxel-based morphometry was done with SPM12 (Wellcome Trust Center for Imaging, London, UK, http://www.fil.ion.ucl.ac.uk/spm), which enables automated spatial normalization, tissue classification and radio-frequency bias correction to be combined with the segmentation step. Cut-off of spatial normalization was 25 mm and medium affine regularization 0.01 was used. Following normalization and segmentation into GM and WM, a modulation step was incorporated to take into account volume changes caused by spatial normalization. Importantly, the modulation step corrects for the differences in total brain size across subjects. Finally, the segmented, normalized, and modulated GM images were smoothed using a Gaussian kernel of 8 mm FWHM.

### Suppelementary Results

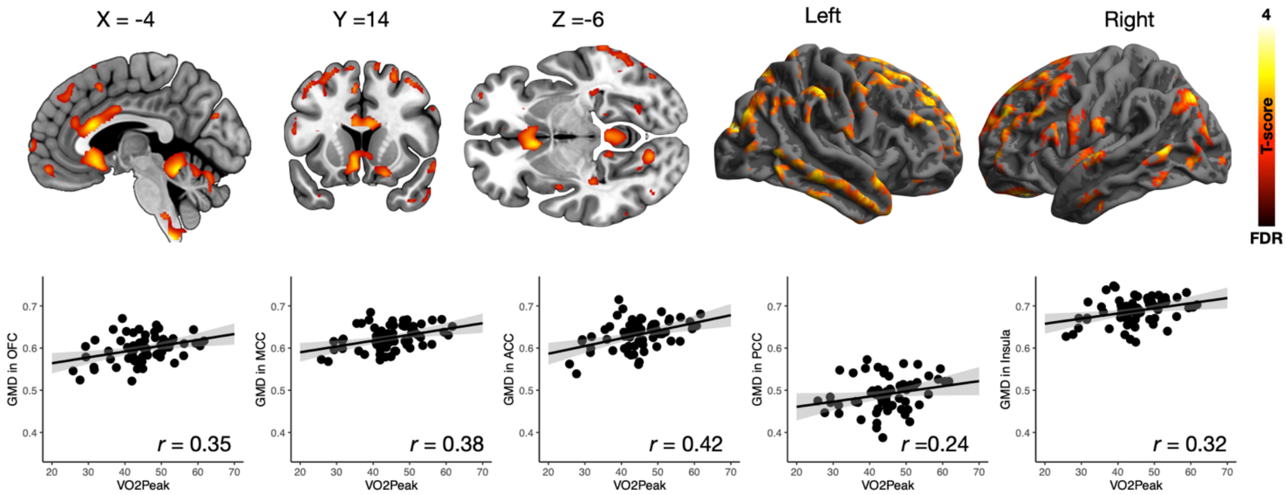


***Figure S1.*** *Positive association between $VO_{2peak}$ and grey matter density. The data are thresholded at $p < 0.05$, FDR-corrected at the cluster level. Scatterplots show the corresponding association (LS-regression line with 95% CI) in representative anatomical regions of interest (GMD; grey matter density).*

**Higher $VO_{2peak}$ and physical activity level are positively associated with GM density**

VBM analyses revealed a positive correlation between $VO_{2peak}$ and GM density when controlling for age and BMI (**Fig. S1**). These associations were found in prefrontal and orbitofrontal cortex, nucleus accumbens, amygdala, primary motor cortex, parietal regions (precuneus, supramarginal gyrus, somatosensory cortex, temporal lobes (superior, middle, and inferior temporal gyri, fusiform gyri, hippocampi), occipital cortex and cerebellum. Self-reported physical activity also correlated positively with GM density in the same regions but only when age was not used as a covariate (data not shown). These effects were most salient in prefrontal regions governing emotional processing and executive functions, limbic regions governing emotional functions, parietal and temporal areas sub-serving long-term memory as well as somatosensory, auditory, speech, and vision processing, and occipital visual cortices, and cerebellum. These regional findings align with those observed in previous studies examining a link between aerobic fitness and cortical structure in older age groups (Erickson *et al*, 2014) and complement the present evidence in younger subjects (Bento-Torres *et al*, 2019; Peters *et al*, 2009; Schlaffke *et al*, 2014; Stillman *et al*, 2018; Whiteman *et al*, 2016): Our data suggest that aerobic fitness might be an important neuroprotective factor already in the early adulthood. The effects of aerobic fitness on MOR availability and GM density were found in opposing directions: Whereas $VO_{2peak}$ was positively associated with GM density, it was negatively associated with MOR availability. Thus, even though good aerobic fitness is in general associated with greater grey matter density, this effect does not pertain to opioidergic neurons, whose density is actually lower in individuals with better fitness. Altogether these data thus suggest that the protective effects of aerobic fitness on CNS are highly specific with respect to the neuroreceptor type; unfortunately, the present single-radioligand study cannot reveal which receptor types show downregulation in sedentary lifestyle.
